# Wake and non-rapid eye movement sleep dysfunction is associated with colonic neuropathology in Parkinson’s disease

**DOI:** 10.1101/2023.10.03.23296499

**Authors:** Mathilde Sadoc, Thomas Clairembault, Emmanuel Coron, Christian Berthomier, Séverine Le Dily, Fabienne Vavasseur, Albane Pavageau, Erik K. St. Louis, Yann Péréon, Michel Neunlist, Pascal Derkinderen, Laurène Leclair-Visonneau

**Author notes:** Corresponding author: Laurène Leclair-Visonneau, Laboratoire d’Explorations fonctionnelles - CHU de Nantes - Bd Jacques Monod 44093 Nantes CEDEX 01 – France. Financial Disclosure/Conflict of Interest: Authors have nothing to disclose regarding the research related to the manuscript.

## Abstract

**Study Objectives:** The body-first Parkinson’s disease (PD) hypothesis suggests initial gut Lewy body pathology that propagates to the pons before reaching the substantia nigra, and subsequently progresses to the diencephalic and cortical levels. This disease course may also be the most likely in PD with rapid eye movement sleep behavior disorder (RBD).

**Objectives:** We aimed to explore the potential association between colonic phosphorylated alpha-synuclein histopathology (PASH) and diencephalic or cortical dysfunction evidenced by non-rapid eye movement (NREM) sleep and wakefulness polysomnographic markers.

**Methods:** In a study involving 43 patients with PD who underwent clinical examination, rectosigmoidoscopy, and polysomnography, we detected PASH on colonic biopsies using whole-mount immunostaining. We performed a visual semi-quantitative and automated quantification of spindle and slow wave features of NREM sleep, and the wake dominant frequency, and then determined Braak and Arizona stage classifications for PD severity based on sleep and wake electroencephalographic features.

**Results:** The visual analysis aligned with the automated quantified spindle characteristics and the wake dominant frequency. Altered NREM sleep and wake parameters correlated with markers of PD severity, colonic PASH, and RBD diagnosis. Colonic PASH frequency also increased in parallel to presumed PD Braak and Arizona stage classifications.

**Conclusions:** Colonic PASH in PD is strongly associated with widespread brain sleep and wake dysfunction, pointing toward likely extensive diffusion of the pathological process in the presumptive body-first PD phenotype. Visual and automated analyses of polysomnography signals provide useful markers to gauge covert brain dysfunction in PD.

**Statement of Significance:** The presence of gut synucleinopathy in Parkinson’s disease can be linked to the body-first hypothesis in its pathophysiology. This study, performed in a cohort of 43 patients with Parkinson’s disease that underwent clinical assessment, rectosigmoidoscopy and polysomnography, provides evidence that colonic neuropathology in Parkinson’s disease is associated with widespread brain dysfunction, as evaluated by wake and non-rapid eye movement sleep polysomnographic markers. Our results support the assumption of an extensive diffusion of the pathological process to diencephalic and neocortical structures in the presumptive body-first phenotype. They also suggest the use of routine polysomnography in phenotyping patients with Parkinson’s disease. Future studies should investigate the brain diffusion pattern and its sleep markers in the hypothesized brain-first phenotype of Parkinson’s disease.

## Introduction

Since first pathological studies published four decades ago, it has been shown that the pathological hallmarks of Parkinson’s disease (PD), namely Lewy bodies and neurites, were found in the gastro-intestinal tract in almost all subjects with PD [1–3]. Digestive phosphorylated alpha-synuclein histopathology (PASH) follows a rostrocaudal gradient of decreasing frequency in PD from lower esophagus to stomach, then ascending colon, descending colon and finally rectum involvement [4,5]. The question of whether enteric neuropathology precedes or occurs in parallel with central nervous system damage remains highly debated [6–8]. Braak and collaborators proposed a well-known staging system for the spread of PD pathology, ranging from stage 1 with lesions limited to the dorsal motor nucleus to stage 6 with diffuse cortical lesions [9]. They also put forward a dual hit hypothesis suggesting that a causal environmental pathogen would penetrate olfactory bulb or enteric nervous system to trigger PD pathology [10]. An alternative staging system developed by the Arizona PD Consortium identified olfactory bulb lesions in stage I, followed by brainstem- or limbic-predominant lesions in stage II, and neocortical involvement in stage IV, whereas peripheral synucleinopathy would not occur until stages II to IV [11]. Considering different clinical phenotypes, Borghammer and collaborators drew parallels between the dichotomy observed in Arizona PD Consortium stage II and clinical phenotypes based on the presence or absence of rapid eye movement sleep behavior disorder (RBD) [12]. They explored the hypothesis of body-first vs brain-first subtypes in Lewy body diseases, in which it is suggested that premotor RBD (i.e. the occurrence of isolated RBD before the motor triad) is a strong predictor of the body-first subtype, as propagating bottom-up pathology will first affect the pons before reaching the substantia nigra [9]. In line with this body-first subtype, we previously showed that colonic synucleinopathy was associated with RBD in subjects with PD [3]. Here, in order to refine and extend these findings, we explored the potential association between colonic PASH and diencephalic or cortical dysfunction, as evaluated by polysomnographic markers, such as spindles and slow waves in non-rapid eye movement (NREM) sleep and the fundamental dominant frequency during wakefulness [13,14].

## Patients and methods

### Participants

The 43 patients with idiopathic PD and exploitable colonic biopsies were derived from a cross-sectional study that took place from February 2013 to February 2016 in Nantes University Hospital, and analyzed the association between RBD and enteric PASH [3], as well as a comprehensive evaluation of autonomic dysfunction in PD [15]. They were prospectively and consecutively recruited according to their disease duration (duration 1 to 5 years: n=14 patients, 5 to 10 years: n=16, longer than 10 years: n=13). Exclusion criteria included confounding factors for autonomic failure or neuropathy, dementia (mini mental status examination MMSE ≤ 23) and irritable bowel syndrome preceding PD. This study was conducted in accordance with the Declaration of Helsinki, approved by the local Ethical Committee (*Comité de protection des personnes Ouest VI, France*) and registered on ClinicalTrials.gov (identifier NCT01748409). All participants provided written informed consent. Collected clinical data included sex, age at onset, disease duration, Unified Parkinson’s Disease Rating Scale part III (UPDRS-III) and its axial subscore, Montreal cognitive assessment (MoCA), MMSE and Parkinson disease sleep scale (PDSS).

### Colonic PASH

As previously described, colonic synucleinopathy assessment was performed on 2 biopsies collected at the junction between sigmoid and descending colon in the course of a rectosigmoidoscopy. Patients with PD were classified as PD+PASH when at least one structure was immunoreactive for both phosphorylated alpha-synuclein and PGP9.5 in whole-mount immunostaining of the submucosa, otherwise classified as PD-PASH [3].

### Polysomnography recordings and visual analysis

A structured interview, sleep questionnaires and overnight video-polysomnography (PSG) were conducted in all 43 patients. PSG recordings were performed using a Natus Deltamed digital system. The montage included bilateral frontal, central, temporal and occipital electroencephalography (EEG) electrodes, bilateral electrooculography, chin and anterior tibialis electromyography. PSG data were interpreted in accordance with the recommendations of the American Academy of Sleep Medicine [16]. Diagnosis of RBD followed the International Classification of Sleep Disorders–II diagnostic criteria [17]. In cases where patients had absent or infrequent (≤ 1%) REM sleep in polysomnographic recording, a diagnosis of probable RBD was considered based on a clinical interview performed by a trained sleep specialist (LLV). Blinded to the patients’ characteristics, visual inspection of N2 and N3 stages in NREM sleep relied on spindles density, slow waves amplitude, and wake rhythm contamination to classify PSG as either normal (numerous spindles per N2 or N3 epochs, high voltage slow wave amplitude, absent or infrequent wake rhythm), severely abnormal (slow < 11 Hz or absent spindles, monotonous and low voltage slow waves, permanent wake rhythm), or moderately abnormal (intermediate recordings that did not meet the criteria for normal or severely abnormal: rare but present spindles – approximately 1/epoch, intermediate voltage or intermittent monotonous pattern, intermittent wake rhythm persistence). The fundamental dominant frequency was derived from the awake resting EEG in the posterior derivations by measuring the dominant frequency during a 15-second period of the most rapid resting wake epoch at the onset of the recording A classification of presumed alpha-synuclein pathologic involvement of the brain, based on the frequency of abnormal wake and sleep electrophysiologic markers (visual NREM sleep characteristics, presence of RBD) was also then applied, according to the pathological classifications proposed by Braak [9] and the Arizona Consortium [11].

### Automated analysis

PSG were also automatically analyzed by using the single-channel ASEEGA algorithm (PHYSIP, Paris, France) which has previously been validated against visual scoring in various sleep centers and conditions: in young [18,19] and older individuals [20], and in clinical settings [21]. The EEG bipolar channel C4-O2 of each of the 43 recordings was analyzed (ASEEGA version R 4.5.20) using the same epoch-splitting as in the visual staging, providing sleep staging together with spindle and slow wave detection and characterization. After automatically adapting the frequency bands based on data-driven criteria intended to account for EEG variability across individuals, NREM sleep features were quantified in N2 + N3 epochs scored unanimously between automatic and visual scorings. Spindles were quantified according to total number and density (number per 30-s epoch). Each detected event was then characterized according to its duration (s), absolute power (µV^2^), purity (ratio sigma power/total power), frequency (mean of instantaneous frequency, Hz), frequential instability (variance of instantaneous frequency, Hz^2^), temporal instability (envelope variations, i.e. quantification of the irregularity of the event shape, µV/s) and maximum amplitude (µV). Slow waves were quantified according to the total number and density (number per 30-s epoch), and each detected event was characterized based on its duration (s), absolute power (µV^2^), temporal instability (oscillations during the slow wave of the delta-filtered signal, µV/s), maximum positive and minimum negative amplitudes as well as peak-to-peak amplitude (µV), first- and second-segment average and maximum slopes, as defined before [22]. Spectral density power was computed for delta, theta, alpha, sigma (spindle frequency range), and beta adapted bands. Automated dominant frequency was computed for each 30-s epoch of wakefulness using an autoregressive model of order 4 after automatic signal cleaning, and defined as the frequency of the maximum of the normalized power spectrum in the [2–14] Hz frequency band.

### Statistics

Clinical, demographic, and PSG data are presented as means and standard deviations. Automatic (AS) and visual (VS) stage scorings were compared on an epoch-by-epoch basis using the percentage agreement, defined as the percentage of epochs that were assigned the same sleep stage, and the Cohen’s kappa coefficient (κ). In addition to the 5-state scoring comparison, AS and VS were compared on the 2-state wake/sleep scoring task as well as in the scoring of the N2-N3 state. Categorical variables were analyzed with Fisher’s exact test or Chi-square when applicable, while continuous variables were analyzed using nonparametric Mann-Whitney U test (when comparing PD+PASH and PD-PASH or PD+RBD+PASH and PD+RBD-PASH groups) or Kruskal-Wallis with Bonferroni post-hoc tests (when comparing 3 groups according to visual classification) using R statistical software (R, Vienna, Austria). Spearman correlation explored associations between PSG and clinical variables (considering non-normally distributed variables). Alpha level was set at p<0.05.

## Results

### Comparison between visual and automated polysomnographic analysis

Among the 43,748 recorded epochs, 1,180 (2.7%) were discarded by AS for signal quality reasons on C4-O2, and 155 (0.4%) by VS, leaving 42,435 epochs for the statistical comparisons. The percentage agreement between AS and VS was 70% with a κ = 0.6, agreement was 90% (κ = 0.8) in sleep/wake scoring and 82% in distinguishing N2 or N3 from other sleep stages. Out of the 43 patients, 15 exhibited normal NREM sleep, 17 had moderately altered NREM sleep and 11 had severely altered NREM sleep based on visual semi-quantitative inspection. To assess the accuracy of visual NREM classification, we compared it with the automated quantification of NREM sleep features: spindles and slow waves characteristics and spectral analysis (Table 1). The frequency of spindles significantly differentiated the three groups, with lower frequencies observed in altered groups. Spindles number and density differentiated normal and moderately altered NREM sleep from severely altered NREM sleep, showing reduced number and density in the latter. Spindles frequential instability was higher in the severely altered NREM sleep group compared to the normal NREM sleep group. Both groups with altered NREM sleep exhibited higher alpha frequency in N2 and N3 stages normalized spectral analysis compared to the normal NREM sleep group. While some variables did not reach statistic differences, several spindles characteristics showed a gradient between the normal group and the severely impaired group, with intermediate values in the moderately impaired group (Table 1). None of the slow wave characteristics differed between the three visually determined groups, although some slope values suggested non-significant gradients. Visually determined fundamental dominant frequency during wakefulness showed a strong correlation with automatically determined dominant frequency (r=0.70, p<0.001).

**Table 1:** Visual and automated NREM sleep analysis comparisons.

|  | Normal NREM sleep (N) | Moderately altered NREM sleep (MA) | Severely altered NREM sleep (SA) | p-value | pairwise comparisons |
| --- | --- | --- | --- | --- | --- |
| number of patients | 15 | 17 | 11 |  |  |
| **Spindles** |  |  |  |  |  |
| Number | 449.1 ± 284.1 | 228.3 ± 116 | 118.4 ± 155.1 | <0.001* | N>SA, MA>SA |
| Density (/30s-epoch) | 1.2 ± 0.6 | 0.7 ± 0.4 | 0.4 ± 0.3 | 0.002* | N>SA, MA>SA |
| Duration (s) | 0.83 ± 0.08 | 0.83 ± 0.06 | 0.80 ± 0.07 | 0.46 |  |
| Power (μV<sup>2</sup>) | 40.1 ± 26.2 | 29.0 ± 23.9 | 22.4 ± 17 | 0.07 |  |
| Purity (%) | 42 ± 7.9 | 36.9 ± 7.7 | 34.9 ± 8 | 0.06 |  |
| Frequency (Hz) | 13.9 ± 0.4 | 13.0 ± 0.7 | 12.5 ± 0.7 | <0.001* | N>MA>SA |
| Frequential instability (Hz<sup>2</sup>) | 0.5 ± 0.1 | 0.6 ± 0.1 | 0.7 ± 0.2 | 0.01* | N<SA |
| Temporal instability (μV/s) | 16.3 ± 13.1 | 14.3 ± 6.5 | 14.4 ± 5.2 | 0.92 |  |
| Maximum amplitude (μV) | 11.8 ± 3.8 | 10 ± 3.9 | 8.8 ± 3.4 | 0.08 |  |
| **Slow waves** |  |  |  |  |  |
| Number | 503.7 ± 459 | 670.8 ± 522.5 | 282.2 ± 255.8 | 0.07 |  |
| Density (/30s-epoch) | 1.5 ± 1.2 | 1.8 ± 1.2 | 1.4 ± 0.9 | 0.42 |  |
| Duration (s) | 1.1 ± 0.1 | 1.1 ± 0.1 | 1.1 ± 0.1 | 0.74 |  |
| Power (μV<sup>2</sup>) | 911.9 ± 541.4 | 662.7 ± 350 | 651.4 ± 426.6 | 0.29 |  |
| Temporal instability (μV/s) | 125.2 ± 74.6 | 97.1 ± 29.8 | 95.2 ± 28.1 | 0.32 |  |
| Maximum positive amplitude (μV) | -49.6 ± 15.6 | -40.8 ± 11.9 | -38.9 ± 14.6 | 0.20 |  |
| Maximum negative amplitude (μV) | 40.3 ± 12.1 | 36.4 ± 10.3 | 37.1 ± 11.1 | 0.60 |  |
| Peak-to-peak amplitude (μV) | 89.9 ± 27.5 | 77.2 ± 22 | 76.1 ± 25.4 | 0.32 |  |
| 1<sup>st</sup> average slope (μV/s) | 273.9 ± 98.3 | 216.8 ± 69.2 | 214.4 ± 82.7 | 0.20 |  |
| 2<sup>nd</sup> average slope (μV/s) | 209.5 ± 71.6 | 185 ± 69 | 165.9 ± 61.2 | 0.18 |  |
| 1<sup>st</sup> maximum slope (μV/s) | 420.1 ± 139.8 | 343.7 ± 103.1 | 336.9 ± 119.8 | 0.25 |  |
| 2<sup>nd</sup> maximum slope (μV/s) | 381.8 ± 120.4 | 347.8 ± 113.1 | 338.1 ± 99.9 | 0.51 |  |
| **NREM sleep spectral density** |  |  |  |  |  |
| Delta band (μV<sup>2</sup>) | 74.3 ± 45.1 | 62.6 ± 38.1 | 58.4 ± 36.3 | 0.63 |  |
| Theta band (μV<sup>2</sup>) | 30.9 ± 20.2 | 30.9 ± 24.7 | 23.1 ± 14.2 | 0.56 |  |
| Alpha band (μV<sup>2</sup>) | 15.1 ± 9.6 | 23.2 ± 19.6 | 29.8 ± 26.9 | 0.44 |  |
| Sigma band (μV<sup>2</sup>) | 7.6 ± 4.1 | 7.0 ± 6.5 | 6.5 ± 5.0 | 0.47 |  |
| Beta band (μV<sup>2</sup>) | 6.2 ± 3.5 | 5.6 ± 6.2 | 5.5 ± 3.0 | 0.41 |  |
| Delta nu band (%) | 0.52 ± 0.07 | 0.49 ± 0.08 | 0.47 ± 0.08 | 0.49 |  |
| Theta nu band (%) | $0.24 \pm 0.04$ | $0.24 \pm 0.05$ | $0.20 \pm 0.04$ | 0.13 | |
| Alpha nu band (%) | $0.12 \pm 0.04$ | $0.18 \pm 0.06$ | $0.22 \pm 0.1$ | 0.003* | N<MA,<br>N<SA |
| Sigma nu band (%) | $0.06 \pm 0.02$ | $0.06 \pm 0.03$ | $0.05 \pm 0.03$ | 0.44 | |
| Beta nu band (%) | $0.06 \pm 0.03$ | $0.05 \pm 0.03$ | $0.05 \pm 0.02$ | 0.09 | |

NREM sleep: non-rapid eye movement sleep, nu: normalized

### Clinical correlates of NREM sleep and wake features

The number and density of spindles showed negative correlations with age, PD duration, UPDRS total and axial scores, and positive correlations with MoCA (except for spindles density), MMSE and PDSS (where a higher score indicates better sleep) (coefficients in Supplementary Table S1). Several spindles alterations correlated with markers of disease severity: duration negatively with UPDRS total score, power negatively with age, frequency negatively with UPDRS axial score and disease duration, frequential instability positively with disease duration, purity negatively with UPDRS total score and age, maximum amplitude negatively with age.

The number of slow waves positively correlated with MoCA, duration positively correlated with age and negatively correlated with disease duration, temporal instability negatively correlated with age, maximum positive amplitude (absolute value) negatively correlated with age, maximum negative amplitude negatively correlated with MMSE. All slow waves slope variables showed a negative correlation with age, and the first average slope showed a correlation with PDSS.

Sigma normalized spectral density negatively correlated with UPDRS total score. PDSS showed a negative correlation with theta, alpha, and beta band absolute spectral density. Manual fundamental dominant frequency positively correlated with MoCA and negatively correlated with UPDRS axial score, while automated dominant frequency negatively correlated with UPDRS axial score and disease duration.

### Clinical, NREM sleep and wake parameters comparisons between PD+PASH and PD-PASH groups

Patients with PD with or without colonic PASH did not differ in terms of age (61.6 ± 6.9 vs 59.8 ± 8.5 years, respectively; p=0.5), sex (15 males / 5 females vs 12/11, respectively; p=0.21), PD duration (123.5 ± 84.4 vs 95.4 ± 75.8 months, respectively; p=0.14) or medications: antidepressants (5/20 vs 3/23, respectively; p=0.44) and benzodiazepines/Z-drugs (6/20 vs 5/23, respectively; p=0.73). Patients with PD+PASH showed a higher frequency of RBD compared to patients with PD-PASH (18/20, 90% vs 10/23, 43.5%, respectively; p=0.002). Colonic PASH was associated with visually altered NREM sleep (moderately or severely altered), lower spindles number, density, power, purity and maximum amplitude, as well as higher frequential instability (Table 2). Slow waves features did not differ between the groups, except for the first maximal slope which was lower in the PD+PASH group. Spectral density analysis revealed lower sigma band power density in the PD+PASH group. The PD+PASH group also exhibited a lower automatically computed dominant frequency during wakefulness and a more frequent visually determined dominant frequency below 8Hz (4/20 20% in PD+PASH vs 0/23 in PD-PASH, p=0.04).

**Table 2:** NREM sleep and wake parameters comparisons between patients with PD, with (PD+PASH) or without colonic PASH (PD-PASH)

|  | PD+PASH | PD-PASH | p-value |
| --- | --- | --- | --- |
| number of patients | 20 | 23 |  |
| Altered visual NREM sleep | 17 (85%) | 11 (47.8%) | 0.02* |
| **Spindles** |  |  |  |
| Number | 364.9 ± 247.9 | 176.4 ± 180 | 0.003* |
| Density (/30s-epoch) | 0.6 ± 0.5 | 0.9 ± 0.5 | 0.02* |
| Duration (s) | 0.8 ± 0.1 | 0.8 ± 0.1 | 0.95 |
| Power ($\mu V^2$) | 21.2 ± 16.8 | 39.0 ± 25.8 | 0.006* |
| Purity (%) | 35.3 ± 7.1 | 40.7 ± 8.5 | 0.03* |
| Frequency (Hz) | 12.9 ± 0.9 | 13.4 ± 0.8 | 0.05 |
| Frequential instability ($Hz^2$) | 0.6 ± 0.1 | 0.5 ± 0.1 | 0.01* |
| Temporal instability ($\mu V/s$) | 15.8 ± 12.2 | 14.4 ± 5.1 | 0.55 |
| Maximum amplitude ($\mu V$) | 8.7 ± 3.2 | 11.6 ± 3.9 | 0.01* |
| **Slow waves** |  |  |  |
| Number | 444.2 ± 349.4 | 573 ± 543 | 0.60 |
| Density (/30s-epoch) | 1.6 ± 0.9 | 1.6 ± 1.3 | 0.64 |
| Duration (s) | 1.1 ± 0.1 | 1.1 ± 0.1 | 0.84 |
| Power ($\mu V^2$) | 603.2 ± 310.9 | 858.1 ± 512.3 | 0.11 |
| Temporal instability ($\mu V/s$) | 91.8 ± 26.7 | 107.2 ± 32.8 | 0.10 |
| Maximum positive amplitude ($\mu V$) | -39 ± 11.5 | -44.3 ± 26.5 | 0.10 |
| Maximum negative amplitude ($\mu V$) | 35 ± 8.6 | 40.3 ± 12.3 | 0.13 |
| Peak-to-peak amplitude ($\mu V$) | 74 ± 19.9 | 87.1 ± 27.6 | 0.12 |
| 1<sup>st</sup> average slope ($\mu V/s$) | 205.2 ± 65.7 | 260 ± 93.7 | 0.07 |
| 2<sup>nd</sup> average slope ($\mu V/s$) | 176 ± 65 | 198.3 ± 71.0 | 0.24 |
| 1<sup>st</sup> maximum slope ($\mu V/s$) | 324.1 ± 97.7 | 403.2 ± 133.6 | 0.047* |
| 2<sup>nd</sup> maximum slope ($\mu V/s$) | 330 ± 105 | 378.6 ± 113.6 | 0.12 |
| **NREM sleep spectral density** |  |  |  |
| Delta band ($\mu V^2$) | 55.1 ± 26.3 | 73.9 ± 47 | 0.25 |
| Theta band ($\mu V^2$) | 23 ± 33.7 | 33.7 ± 24.7 | 0.13 |
| Alpha band ($\mu V^2$) | 19.9 ± 18.8 | 24.1 ± 20.7 | 0.47 |
| Sigma band ($\mu V^2$) | 5.4 ± 5.1 | 8.4 ± 5.2 | 0.02* |
| Beta band ($\mu V^2$) | 4.5 ± 3.2 | 6.8 ± 5.3 | 0.11 |
| Delta nu band (%) | 0.49 ± 0.1 | 0.50 ± 0.08 | 0.73 |
| Theta nu band (%) | 0.22 ± 0.05 | 0.23 ± 0.05 | 0.64 |
| Alpha nu band (%) | 0.17 ± 0.07 | 0.16 ± 0.08 | 0.54 |
| Sigma nu band (%) | 0.05 ± 0.02 | 0.07 ± 0.03 | 0.13 |
| Beta nu band (%) | 0.05 ± 0.02 | 0.05 ± 0.02 | 0.78 |
| **Wake parameters** |  |  |  |
| Manual dominant frequency (Hz) | 8.9 ± 1.3 | 9.6 ± 1.1 | 0.07 |
| Automated dominant frequency (Hz) | 8.6 ± 0.8 | 9.2 ± 0.9 | 0.03* |

nu: normalized, PASH: phosphorylated alpha-synuclein histopathology in colonic biopsies,
PD: Parkinson disease

Considering the covariate of RBD, we conducted the same inter-group comparisons based on RBD status. Among the 15 patients without RBD, 2 had PD-RBD+PASH while 13 patients had PD-RBD-PASH. There was no difference in visually altered NREM sleep frequency between patients with PD-RBD (1/2, 50% in PD-RBD+PASH vs 5/13, 38.5% in PD-RBD-PASH, p=1). These relatively small sample sizes for PD-RBD patients precluded any informative statistical analysis for quantitative variables between these subgroups with and without PASH positive pathology. Among the 28 patients having PD with RBD, 18 had positive colonic PASH while 10 did not. Patients with PD+RBD with or without colonic PASH did not differ in age (62.6 ± 6.1 vs 61.8 ± 7.2 years, respectively; p=0.56), sex (14/4 vs 5/5, respectively; p=0.21), PD duration (127.9 ± 87.3 vs 113.1 ± 74.6 months, respectively; p=0.50) or medications: antidepressants (5/18 vs 0/10, respectively; p=0.13) and benzodiazepines/Z-drugs (5/18 vs 3/10, respectively; p=0.1). Patients with PD+RBD+PASH showed fewer spindles (in terms of number and density), lower spindles power, purity, and maximum amplitude compared to patients with PD+RBD-PASH (Table 3). Slow waves in the PD+RBD+PASH group showed lower first average slope and first maximal slope, while temporal instability was reduced. Spectral density analysis revealed lower sigma (absolute and normalized) and beta band power density in patients with PD+RBD+PASH compared to those with PD+RBD-PASH. Wake fundamental dominant frequencies did not differ between the groups, and 4 patients in the PD+RBD+PASH group had visually determined dominant frequency below 8Hz (4/18, 22%) compared to none in the PD+RBD-PASH group (p=0.27).

**Table 3:** NREM sleep and wake parameters comparisons between patients with PD and RBD, with (PD+RBD+PASH) or without colonic PASH (PD+RBD-PASH)

|  | PD+RBD+PASH | PD+RBD-PASH | p-value |
| --- | --- | --- | --- |
| number of patients | 18 | 10 |  |
| Altered visual NREM sleep | 16 (89%) | 6 (60%) | 0.15 |
| **Spindles** |  |  |  |
| Number | 143.8 ± 114.8 | 297.1 ± 230.7 | 0.03* |
| Density (/30s-epoch) | 0.5 ± 0.4 | 0.8 ± 0.4 | 0.04* |
| Duration (s) | 0.8 ± 0.1 | 0.8 ± 0.1 | 0.94 |
| Power ($\mu V^2$) | 17.4 ± 12.2 | 45.3 ± 31.1 | 0.002* |
| Purity (%) | 34.0 ± 6.0 | 43.1 ± 9.6 | 0.007* |
| Frequency (Hz) | 12.9 ± 0.9 | 13.5 ± 0.8 | 0.06 |
| Frequential instability ($Hz^2$) | 0.5 ± 0.1 | 0.6 ± 0.2 | 0.12 |
| Temporal instability ($\mu V/s$) | 15.1 ± 12.5 | 15.6 ± 5.4 | 0.19 |
| Maximum amplitude ($\mu V$) | 8 ± 2.5 | 12.5 ± 4.3 | 0.003* |
| **Slow waves** |  |  |  |
| Number | 453.6 ± 350.1 | 329.2 ± 266.5 | 0.41 |
| Density (/30s-epoch) | 1.6 ± 0.9 | 1.1 ± 0.9 | 0.07 |
| Duration (s) | 1.1 ± 0.1 | 1.0 ± 0.1 | 0.24 |
| Power ($\mu V^2$) | 566.1 ± 281.5 | 859.8 ± 581.6 | 0.16 |
| Temporal instability ($\mu V/s$) | 88.1 ± 23.6 | 110.1 ± 34 | 0.002* |
| Maximum positive amplitude ($\mu V$) | -37.7 ± 11 | -46.7 ± 16.2 | 0.14 |
| Maximum negative amplitude ($\mu V$) | 34.1 ± 8 | 40.8 ± 13.5 | 0.22 |
| Peak-to-peak amplitude ($\mu V$) | 71.8 ± 18.8 | 87.5 ± 29.2 | 0.17 |
| 1<sup>st</sup> average slope ($\mu V/s$) | 196.5 ± 61.8 | 273.9 ± 104.9 | 0.046* |
| 2<sup>nd</sup> average slope ($\mu V/s$) | 165.1 ± 52.9 | 198.8 ± 76.7 | 0.20 |
| 1<sup>st</sup> maximum slope ($\mu V/s$) | 311.3 ± 91.1 | 418 ± 146.7 | 0.045* |
| 2<sup>nd</sup> maximum slope ($\mu V/s$) | 314.3 ± 88.8 | 390.7 ± 115.4 | 0.08 |
| **NREM sleep spectral density** |  |  |  |
| Delta band ($\mu V^2$) | 52.6 ± 26 | 67.9 ± 45.5 | 0.50 |
| Theta band ($\mu V^2$) | 21.5 ± 11.1 | 29.7 ± 17.9 | 0.20 |
| Alpha band ($\mu V^2$) | 17.2 ± 14.8 | 21.4 ± 17.5 | 0.24 |
| Sigma band ($\mu V^2$) | 4.3 ± 2.8 | 9.2 ± 5.7 | 0.003* |
| Beta band ($\mu V^2$) | 4.1 ± 2.9 | 8.9 ± 6.9 | 0.01* |
| Delta nu band (%) | 0.51 ± 0.07 | 0.47 ± 0.13 | 0.69 |
| Theta nu band (%) | 0.23 ± 0.05 | 0.22 ± 0.04 | 0.65 |
| Alpha nu band (%) | 0.17 ± 0.07 | 0.16 ± 0.08 | 0.65 |
| Sigma nu band (%) | 0.05 ± 0.02 | 0.07 ± 0.03 | 0.01* |
| Beta nu band (%) | 0.05 ± 0.02 | 0.07 ± 0.03 | 0.13 |
| **Wake parameters** |  |  |  |
| Manual dominant frequency (Hz) | 8.8 ± 1.2 | 9.4 ± 0.9 | 0.16 |
| Automated dominant frequency (Hz) | 8.6 ± 0.8 | 9.2 ± 0.9 | 0.09 |

nu: normalized, PASH: phosphorylated alpha-synuclein histopathology in colonic biopsies,
PD: Parkinson disease, RBD: rapid eye movement sleep behavior disorder

### Colonic PASH following PD stages

When considering either Braak [9] or the Arizona Consortium [11] PD staging, the progression of the PD pathological process reaches the locus subcoeruleus early, followed by diencephalic structures, and eventually the neocortex at a later stage. In a previous study, we found that colonic PASH occurred in 64.3% of patients having PD with RBD compared to 13% in patients with PD without RBD [3]. Among those with RBD, colonic PASH was observed in 72.7% of patients with visually altered NREM sleep, while it was present in 16.7% of those with normal NREM sleep. In patients with PD without RBD, colonic PASH occurred in 33.3% of those with visually altered NREM sleep, while in 11.1% of those with normal NREM sleep. Among the 43 patients with PD, four individuals had visually determined abnormal wake dominant frequency (below 8 Hz), all of whom exhibited RBD, visually altered NREM sleep, and colonic PASH. In case of RBD and altered NREM sleep with a dominant frequency in wake above 8 Hz, colonic PASH was observed in 66.7% of patients with PD.

The frequency of colonic PASH, based on the degree of brain dysfunction determined through visual analysis of PSG (presence of RBD, altered NREM sleep, and abnormal slowing during wakefulness) and categorized according to the PD stages, is illustrated in the Figure.

## Discussion

In the present study, we sought to assess diencephalic and neocortical functions by visually analyzing NREM sleep and wake features in patients with PD, along with assessment for the presence or absence of enteric synucleinopathy pathology. Our semi-quantitative NREM sleep visual analysis aligned with the automated quantification of several spindle characteristics, but was less effective in discriminating slow wave parameters, suggesting that the visual analysis primarily relied on spindles properties. The visually determined dominant frequency during the most rapid wake epoch showed a strong correlation with the automated determination over the total wake time in PSG. Alterations in NREM sleep and wake parameters were correlated with markers of PD severity. When comparing patients with PD with or without colonic PASH, we observed that the changes in NREM sleep and wakefulness were linked to presence of the pathological process within the enteric nervous system, which was also evident in patients with PD and RBD. Last, we estimated the frequency of colonic PASH in our cohort based on the sleep and wake analysis of brain function, revealing an increase in the colonic pathological burden as the PD stages progressed.

NREM sleep spindles are rapid oscillations (11-16 Hz) that arise from thalamo-cortical circuits involving thalamo-cortical and reticular neurons in the thalamus and cortical regions [13,23]. These spindles, together with hippocampal ripples and cortical slow waves, are believed to play a role in memory consolidation through hippocampo-cortical coordination [24]. Previous studies have demonstrated a reduction in spindle density in patients with PD compared to controls, and this reduction has been associated with an increased risk of developing dementia over a mean follow-up of 4.5 years [25,26]. In our cohort, we confirmed correlations between spindle characteristics and cognitive decline, and we additionally showed correlations with motor severity, including axial impairment. Patients with PD and colonic PASH presented with greater alterations in spindles compared to the PD-PASH group. Given that spindles result from thalamo-cortical interactions, spindle alterations might reflect damage to either the thalamus, cortex, or both. NREM sleep slow waves arise from alternating depolarization (up-state) and hyperpolarization (down-state) phases of cortical neurons [13]. Reduction of slow wave amplitude in NREM sleep have been reported in PD, but no association with dementia-risk was found [26]. However, the percentage of N3 stage of NREM sleep has been correlated with global cognition and executive function, language, and processing speed [27]. Some studies suggest that frontal delta slow waves may contribute to cognitive impairment in PD [27,28]. Additionally, a retrospective study found that higher accumulated power of slow waves in NREM sleep was associated with slower motor progression, particularly in axial motor symptoms, in PD [29]. In our present study, we found correlations between slow waves number, negative amplitude and cognitive tests, but only slight differences in slow wave features were observed between patients with PD with or without PASH, suggesting that slow waves may have only a weak association with colonic synucleinopathy. Therefore, it appears that alterations in spindles are more likely attributable to thalamus dysfunction than cortical involvement. Nevertheless, we also showed some cortical involvement associated with enteric synucleinopathy in the form of lower wake dominant frequency.

How do these results relate to the body-first vs brain-first theory of PD pathophysiology [12]? In our previous study with the same cohort, we demonstrated an association between RBD and colonic synucleinopathy [3]. In the current study, we focused on a later stage of PD, specifically stage 4 according to Braak staging or stage III according to the Arizona Consortium, with regard to body diffusion [9,11]. Among the 20 patients with PD+PASH, 90% also had RBD and exhibited greater impairment in spindles, suggesting strong overlap between dorsal pontine and diencephalic/thalamic dysfunction that most likely followed a body-first trajectory. Interestingly, all four patients with visually determined wake frequency below 8Hz displayed colonic synucleinopathy, suggesting a diffuse phenotype in which body-first and brain-first trajectories may converge. However, while the PD+PASH group showed lower occipital wake frequencies indicative of cortical dysfunction, both the PD with or without PASH groups did not differ in terms of age or disease duration, contradicting the notion of late brain involvement from an evolving body-first trajectory in patients with presumptive primary colonic pathology. Furthermore, none of our patient in the PD-PASH group exhibited visually abnormal wake fundamental frequency, suggesting that none presented with a brain-first late stage. Taken together, our findings would support a body-first and diffuse phenotype of PD, with greater involvement of the diencephalon and cortex in patients with PD+RBD+PASH, while patients without PASH may instead exhibit a brain-first and restricted phenotype.

Polysomnography is a standard examination in synucleinopathies, for the accurate diagnosis of RBD and for assessing nocturnal sleep and sleepiness throughout the course of Lewy body disease progression from its prodromal stages to eventual manifest PD [30,31]. We developed a visual and straightforward analysis with semi-quantification of spindles that led to informative NREM sleep classification that was strongly associated with colonic pathology and RBD diagnosis. This classification of NREM sleep, as either altered or normal, can easily be reproduced by sleep specialists in neurology during the routine interpretation of polysomnograms in patients with PD, thereby assessing the probable pathologic diffusion and phenotype of PD. Reduced spindle density and altered thalamic resting state functional connectivity have been observed in isolated RBD [25,32,33]. Furthermore, impaired temporal coupling between slow oscillations and spindles, as well as impaired thalamic resting-state functional connectivity, have been correlated with cognitive decline [33,34]. While changes of cyclic alternating pattern in NREM sleep have been predictive of phenoconversion [35], the predictive value of spindle alteration for the phenoconversion risk of isolated RBD remains to be determined. Impairment in wakefulness EEG, particularly in occipital dominant frequency, has also shown predictive value in assessing the risk of phenoconversion [36]. Further research on our simplified classification of NREM sleep could be conducted in prodromal populations to evaluate its predictive value.

Our study has several limitations. First, the sample size was relatively small, which may have resulted in insufficient statistical power to discriminate groups using certain variables, especially when comparing PD+RBD with or without PASH groups. Additionally, the analysis of colonic PASH was performed with whole-mount immunostaining, which may be less sensitive compared to more advanced and recent techniques such as real-time quaking-induced conversion (RT-QuIC) and protein misfolding cyclic amplification (PMCA) assays for alpha-synuclein detection [37], which should be incorporated into future studies. Of note, the percentage agreement between automated and visual scoring for sleep stages was below the agreement usually observed with patients diagnosed with various sleep disorders; however, this result may reflect the sleep dissociation, especially observed in synucleinopathies.

In conclusion, our results provide evidence for an association between colonic spreading of PD and widespread brain dysfunctions involving diencephalic and cortical structures.

Additionally, our study highlights the potential utility of routine polysomnographic assessment in evaluating the diffusion pattern of the disease in patients with PD. Future research could focus on investigating the brain diffusion pattern and its polysomnographic markers in the hypothesized brain-first phenotype of PD [12], while our study emphasizes the extensive diffusion of the pathological process in patients with a body-first phenotype.

## Data Availability

All data produced in the present study are available upon reasonable request to the authors.

## Acknowledgment

The authors thank the patients for their participation in the study.

---

**Figure.**
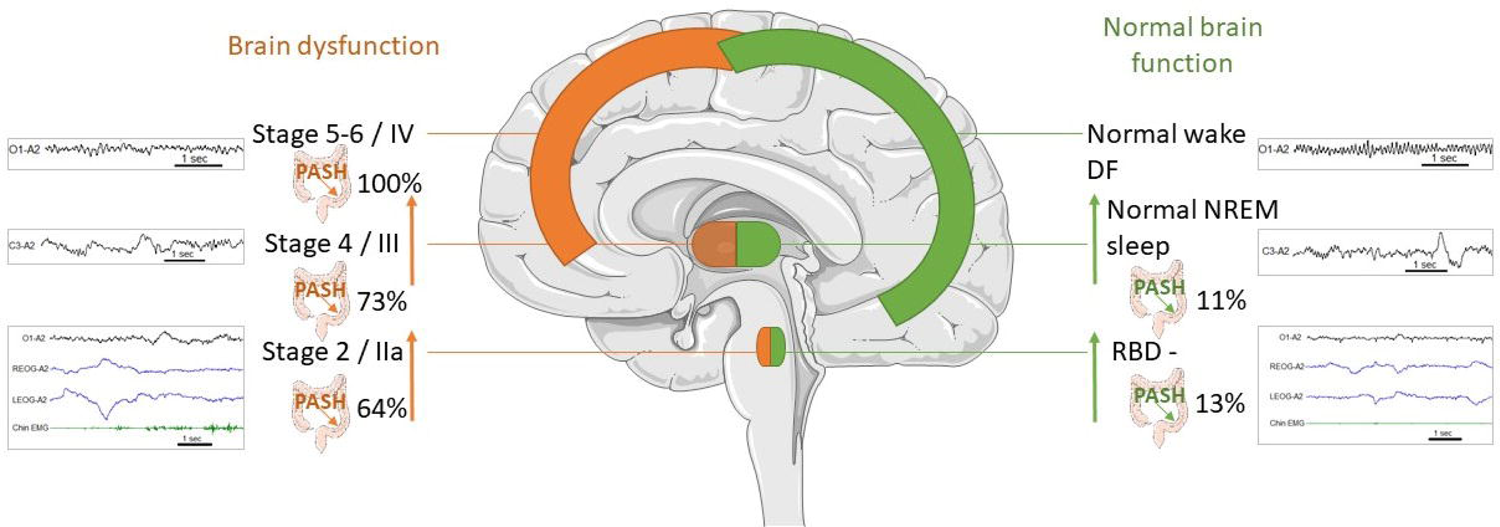
Frequency of colonic PASH based on the brain functioning (all normal or all altered functions: RBD, NREM sleep and wake DF with polysomnogram illustrations) through visual analysis of PSG and categorized according to the PD stages (Braak [9]/Arizona Consortium [11]). Colonic PASH occurred in 64% of patients with PD and RBD (PD+RBD), in 73% of patients with PD+RBD+altered NREM sleep (stage 4/III), and in 100% of patients with PD+RBD+altered NREM sleep+altered wake DF (stage 5-6/IV). Colonic PASH occurred in 13% of patients with PD without RBD (PD-RBD), and in 11% of patients with PD-RBD+normal NREM sleep, all of them had normal wake DF. DF: dominant frequency, EMG: electromyogram, LEOG: left electrooculogram, NREM: non-rapid eye movement, PASH: phosphorylated alpha-synuclein histopathology in colonic biopsies, RBD-: absence of rapid eye movement sleep behavior disorder, REOG: right electrooculogram. C3-A2 and O1-A2 refer to electroencephalography leads.

**Table S1:** Clinical correlates of NREM sleep and wake parameters (Spearman coefficients)

|  | age | PD duration | UPDRS-III total | UPDRS-III axial | MoCA | MMSE | PDSS |
| --- | --- | --- | --- | --- | --- | --- | --- |
| **Spindles** |  |  |  |  |  |  |  |
| Number | -0.37* | -0.44** | -0.54*** | -0.46** | 0.31* | 0.39** | 0.35* |
| Density | -0.35* | -0.32* | -0.61*** | -0.46** |  | 0.40** | 0.33* |
| Duration |  |  | -0.45** |  |  |  |  |
| Power | -0.34* |  |  |  |  |  |  |
| Frequency |  | -0.31* |  | -0.31* |  |  |  |
| Frequential instability |  | 0.43** |  |  |  |  |  |
| Purity | -0.33* |  | -0.37* |  |  |  |  |
| Maximum amplitude | -0.33* |  |  |  |  |  |  |
| **Slow waves** |  |  |  |  |  |  |  |
| Number |  |  |  |  | 0.35* |  |  |
| Duration | 0.31* | -0.36* |  |  |  |  |  |
| Temporal instability | -0.32* |  |  |  |  |  |  |
| Maximum positive amplitude | -0.32* |  |  |  |  |  |  |
| Maximum negative amplitude |  |  |  |  |  | -0.31* |  |
| 1<sup>st</sup> average slope | -0.37* |  |  |  |  |  |  |
| 2<sup>nd</sup> average slope | -0.37* |  |  |  |  |  |  |
| 1<sup>st</sup> maximum slope | -0.33* |  |  |  |  |  |  |
| 2<sup>nd</sup> maximum slope | -0.37* |  |  |  |  |  |  |
| **NREM sleep spectral density** |  |  |  |  |  |  |  |
| Sigma nu band |  |  | -0.34* |  |  |  |  |
| Theta band |  |  |  |  |  |  | -0.33* |
| Alpha band |  |  |  |  |  |  | -0.32* |
| Beta band |  |  |  |  |  |  | -0.35* |
| **Wake parameters** |  |  |  |  |  |  |  |
| Manual dominant frequency |  |  |  | -0.34* | 0.31* |  |  |
| Automated dominant frequency |  | -0.39* |  | -0.32* |  |  |  |

p-values: 0.05>★>0.01>★★>0.001>★★★
MMSE: mini mental state examination, MoCA: Montreal cognitive assessment, PD:
Parkinson disease, PDSS: Parkinson disease sleep scale, UPDRS: Unified Parkinson's
Disease Rating Scale

